# Functional connectome mediates the association between sleep disturbance and mental health in preadolescence

**DOI:** 10.1101/2021.08.12.21261990

**Authors:** Fan Nils Yang, Tina Tong Liu, Ze Wang

**Affiliations:** Department of Diagnostic Radiology and Nuclear Medicine, University of Maryland School of Medicine, Baltimore, MD, USA; Section on Neurocircuitry, Laboratory of Brain and Cognition, National Institute of Mental Health, National Institutes of Health, Bethesda, MD, USA

**Keywords:** functional neuroimaging, sleep, adolescent psychiatry, cognition

## Abstract

**Background:** Sleep disturbance is known to be associated with various mental disorders and typically precedes the onset of mental disorders in youth. Given the increasingly acknowledged bi-directional influence between sleep disturbance and mental disorders, we addressed the possibility of a shared neural mechanism that underlies sleep disturbance and mental disorders in preadolescents.

**Methods:** We analyzed a dataset of 9350 9-10 year-old children, among whom 8845 had one-year follow-up data, from the Adolescent Brain and Cognitive Development (ABCD) Study. Linear mixed-effects models, mediation analysis, and longitudinal mediation analysis were used.

**Results:** Out of 338 unique connectivities, the effect of total sleep disturbance (TSP, from Sleep Disturbance Scale) and total problems (TP, from Child Behavior Checklist) converged in the default mode network (DMN) and its anti-correlated dorsal attention network (DAN). Within- and between-network connectivities (DMN-DAN, DMN-DMN, DAN-DAN) mediated the relationship between baseline TSD and TP at one-year follow-up and the relationship between baseline TP and TSD at one-year follow-up. Moreover, the identified network connectivities (DMN-DAN, DAN-DAN) also correlated with the total cognitive composite score from the NIH toolbox.

**Conclusions:** The pathway model in which sleep disturbance and mental problems affect each other through two anticorrelated brain networks (DMN and DAN) suggests a common neural mechanism between sleep disturbance and mental disorders. A less segregated DMN and DAN is also associated with negative outcomes on mental well-being, sleep disturbance, and cognition. These findings have important implications for the design of prevention and neurofeedback intervention for mental disorders and sleep problems.

**Highlights:**

1. The impact of sleep disturbance and mental disorders on functional connectivity converged in default mode and dorsal attention networks.
2. The relationship between sleep disturbance and mental disorders was mediated via a shared brain network mechanism
3. Sleep disturbance and mental disorders at baseline can each predict the other one year later through the identified network connectivities
4. A less segregated default mode and dorsal attention networks was correlated with poorer cognitive performance.

## Introduction

Mental disorders tend to share similar risk factors or biomarkers, and respond to the same therapies (Caspi and Moffitt, 2018). Both theoretical and empirical evidence suggests that this comorbidity might be due to the nonspecificity of the functional neurocircuit (Lees et al., 2021; McTeague et al., 2017; Menon, 2011; Sha et al., 2019). Specifically, according to the triple network model, a wide variety of psychopathologies are associated with aberrant functional connectivity within and between three large-scale neurocognitive networks, i.e. salience network, frontoparietal network, and default mode network, as well as subnetworks such as ventral and dorsal attention networks (Menon, 2011). This triple network model is further corroborated by recent results (Lees et al., 2021; McTeague et al., 2017; Sha et al., 2019), including one study that demonstrated that common functional network disruptions in default mode, ventral and dorsal attention networks were linked to all dimensions of psychopathology in preadolescents (Lees et al., 2021).

Moreover, individuals with mental disorders often experience varying degrees of sleep disturbance (Baglioni et al., 2016; Tesler et al., 2013). Recent studies showed that sleep disturbance and mental disorders could aggravate each other in a reciprocal manner (Alfano et al., 2007; Cox and Olatunji, 2016; Gregory and Sadeh, 2016; Hansen et al., 2014; Tesler et al., 2013). On the one hand, longitudinal studies have shown that sleep disturbance in youth likely precedes and exacerbates symptoms of attention deficit hyperactivity disorder (ADHD) (Scott et al., 2013) and mood and anxiety disorders (Goldstone et al., 2020; Gregory et al., 2009, 2005; Jansen et al., 2011). On the other hand, greater polygenic risk factors for various mental disorders, i.e. ADHD, mood and anxiety disorders, may contribute to greater sleep disturbance among children (Ohi et al., 2021). These findings strongly suggest a potentially shared neural mechanism between sleep disturbance and mental disorders, which has not been demonstrated to date.

The purpose of this study was to address this open question using resting-state functional connectivity (rs-FC). rs-FC is an emerging fMRI technique for studying brain networks and cognition as well as their alterations related to brain disease (Gabrieli et al., 2015; Woo et al., 2017). rs-FC measure is highly reproducible within an individual across scan sessions (Finn et al., 2015; Noble et al., 2019). Thus, it has been recognized as a promising neural biomarker for assessing neurocognitive development and identifying aspects of intrinsic brain network organization that are related to cognition and disease status. Similar to the role of the triple network model in mental disorders, altered functional connectivity within and between the default mode network and its anticorrelated networks, including dorsal and ventral attention networks, were found in sleep-deprived adults (Chee and Zhou, 2019; De Havas et al., 2012; Kaufmann et al., 2016; Sämann et al., 2010).

However, few studies have investigated the associations between functional connectivity and sleep disturbance in preadolescence, which is a critical period for brain development. Coincidentally, the onset of mental disorders often starts around the same time, i.e. childhood or adolescence (Kessler et al., 2007; Tesler et al., 2013). To fill the gap of associations between sleep disturbance, mental disorders, and functional connectivity in preadolescence, we analyzed the large data from the Adolescent Brain Cognitive Development (ABCD, https://abcdstudy.org) study, the largest observational and normative project (over 11,000 children) on brain development and child health to date.

We hypothesized that sleep disturbance and mental disorders have similar effects on rs-FC. Out of 338 unique connectivity measures tested, the effect of sleep disturbance and mental disorders converged in the between- and within-network connectivities in the default mode network and one of its anti-correlated networks (i.e. dorsal attention network). In addition to identifying network connectivities that mediated the association between sleep disturbance and mental disorders, we revealed the associations between network connectivities and cognition. Longitudinal analyses demonstrated that these three network connectivities mediated the effect of sleep disturbance on mental disorders one year later and the effect of mental disorders on sleep disturbance one year later.

## Methods

### Participants

The ABCD study includes baseline data from more than 11,000 9-10 year olds (Casey et al., 2018). Data were collected from 21 sites across the United State, approved by institutional review boards (IRB) at the University of California, San Diego as well as at each local site. Parents’ written informed consent and children’s assent were obtained at each site. Recruitment followed demographic distribution (sex, race, ethnicity, household income, etc.) of the general population in the United States. Details about the protocols are available at the ABCD study website (https://abcdstudy.org/scientists/protocols/).

In total, data from 11,878 children are provided at baseline. Among them, 9387 passed the rsfmri QC (imgincl_rsfmri_include) provided by ABCD study (Hagler et al., 2019). An additional 37 children were excluded due to missing values on network connectivity (n=14), total problems (n=21), or total sleep disturbance (n=2). Thus, the total number of children included in the current study was 9350. Table 1 provides the detailed demographic information.

**Table 1.** Demographic information of the participants from the ABCD dataset included in the present study.

| Characteristic | Baseline | 1 year Follow-up |
| --- | --- | --- |
|  | N or Mean (SD) | N or Mean (SD) |
| Age (month) | 119.3 (7.5) | 131.4 (7.8) |
| Sex at birth |  |  |
| female | 4674 | 4412 |
| male | 4676 | 4433 |
| Race |  |  |
| White | 6101 | 5872 |
| Black | 1340 | 1198 |
| Other (mixes or Asians) | 1909 | 1775 |
| Pubertal Status | 1.8 (0.9)<sup>1</sup> | 2.1 (1.0)<sup>3</sup> |
| Total Problems | 45.4 (11.2) | 45.0 (11.0)<sup>4</sup> |
| Total Sleep Disturbance | 36.3 (8.0) | 36.5 (7.9)<sup>5</sup> |
| Total Cognitive Composite Score | 48.3 (11.1)<sup>2</sup> | N/A<sup>6</sup> |
| Number of children | 9350 | 8845 |

Note:
1. 305 children have missing values; 2. 752 children have missing values; 3. 3090 children have missing values; 4. 33 children have missing values; 5. 28 children have missing values; 6. Total cognitive composite score did not have 1 year follow-up data.

### Sleep measures

Total sleep disturbance (TSD) and its one year follow-up was calculated from the ABCD Parent-reported Sleep Disturbance Scale for Children (abcd_sdss01) as the sum of six scores corresponding to six different sleep disorders, including Disorders of Initiating and Maintaining Sleep, Sleep Breathing disorders, Disorder of Arousal, Sleep-Wake transition Disorders, Disorders of Excessive Somnolence, and Sleep Hyperhidrosis (Bruni et al., 1996).

### Mental health measures

Children’s dimensional psychopathology and adaptive functioning were assessed by the Parent-reported Child Behavior Checklist Scores (abcd_cbcls01). The total score of psychiatric problems (total problems t score) and its one year follow-up was calculated based on scores of ten empirically-based syndromes, i.e. anxious/depressed, withdrawn/depressed, somatic complaints, social problems, thought problems, attention problems, rule-breaking behavior, aggressive behavior, internalizing broad band score and externalizing broad band score (Achenbach and Rescorla, 2004).

### Cognitive measures

Total Cognitive Composite Fully-Corrected T-score (abcd_tbss01, baseline only, not data available at one year follow-up) of the NIH Cognition Battery Toolbox was used as the measure for general cognitive function measure (Akshoomoff et al., 2013). It is the sum of the scores from seven cognitive components: language vocabulary knowledge, attention, cognitive control, working memory, executive function, episodic memory, and language.

### Network connectivity

Preprocessing of the functional connectivity analysis and the network connectivity strengths were provided by the ABCD consortium. Detailed MRI scan parameters and preprocessing steps have been reported and discussed (Hagler et al., 2019). Briefly, preprocessing of rs-FC included registration, distortion correction, normalization, regression of 24 motion parameters, outliers with framewise displacement higher than 3mm, and signals from whither matter, cerebral spinal fluid, and whole-brain. Within- or between-network connectivity were calculated as the average fisher-transferred functional connectivity between each pair of ROIs within or between networks (13 networks in total) based on the Gordon altas (Gordon et al., 2016). In addition, network connectivity between each network and each subcortical region (19 subcortical regions in total) were also calculated (Fischl et al., 2002). Quality controls were performed by trained ABCD staff (Hagler et al., 2019) and were used as an inclusion criterion. Sites effects were harmonized by the ComBat method (Johnson et al., 2007; Yang et al., 2021; Yu et al., 2018). Network connectivity measures were not available at one year follow-up.

### Statistical analyses

Linear Mixed-effects Models (LME, implemented through function *fitlme* in Matlab) were used to investigate the effect of total problems or total sleep disturbance on network connectivity. All models included fixed-effect covariates for age, sex at birth, race (black, white, and others), pubertal status (1-4, assessed by ABCD Youth Pubertal Development Scale and Menstrual Cycle Survey History), and random effects for family relatives nested within sites.

The mediation toolbox (https://github.com/canlab/MediationToolbox) was used to perform all the mediation analyses (Wager et al., 2009, 2008). Here, the independent variable is total sleep disturbance/total problems, the dependent variable is total problems/total sleep disturbance, and the mediator is the network connectivity identified through LME. All above-mentioned covariates (age, sex, race, pubertal status, family relatives, and sites) were controlled in the mediation analyses. The significance of the mediation analyses was estimated using bootstrap sampling with 10,000 random-generated samples.

### Longitudinal analyses

8845 of the 9350 children participated in the one-year follow-up study (see Table 1). Mediation analyses were performed to test the longitudinal associations between total sleep disturbance, total problems, and network connectivities. Specifically, we tested whether network connectivities mediated the associations between total sleep disturbance at baseline and total problems one year later (controlled for baseline total problems), and between total problems at baseline and total sleep disturbance one year later (controlled for baseline total sleep disturbance). Additional covariates including baseline age, sex, race, baseline pubertal statuse, familiy relatives, and sites.

## Results

Total sleep disturbance and network connectivity

The distribution of TSD was skewed, so we log-transformed the data to approximate normality. In total, 91 within- and between-network connectivities and 247 network-subcortical connectivities were tested (Fig. 1). Out of the 338 unique comparisons, TSD had a significant impact on only three network connectivities (FDR corrected for multiple comparisons, see asterisks in Fig. 1), DMN-DAN (*β* = 0.0329, *R*^*2*^ = 0.0638, *p* < 1e-4), DMN-DMN (*β* = -0.0326, *R*^*2*^ = 0.0489, *p* = 0.0003, and DAN-DAN (*β* = -0.0355, *R*^*2*^ = 0.0241, *p* = 0.0003).

**Figure 1.**
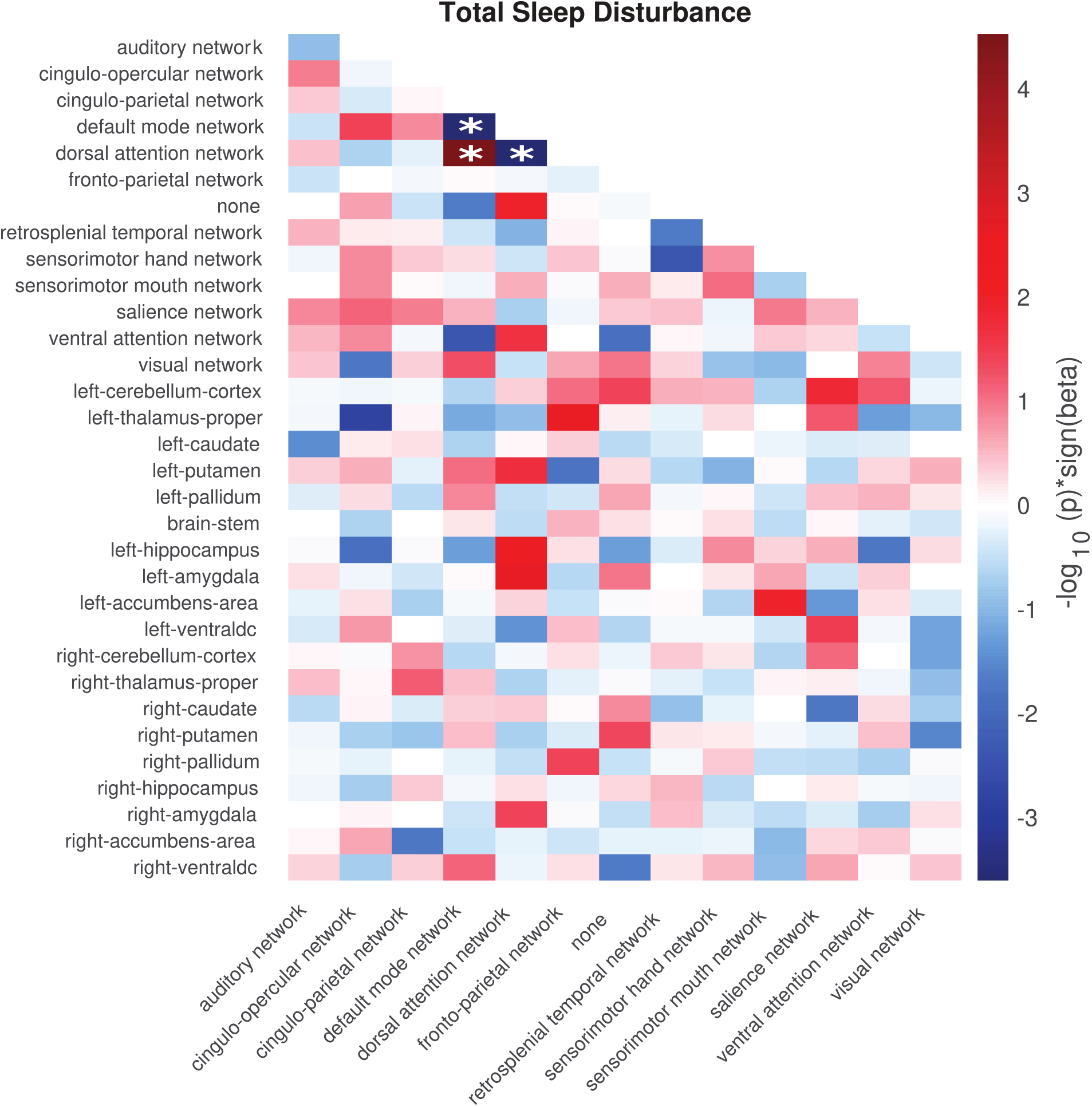
The effect of total sleep disturbance on network connectivity measures. Color represents negative log-transformed *p* value times the sign of beta value of total sleep disturbance. That is, red/blue means positive/negative association between total sleep disturbance and network connectivity measures, respectively. Only unique network connectivity measures were shown in the matrix (i.e. top right was intentionally left blank). * denotes network connectivity that survived FDR correction for multiple comparisons out of 338 unique comparisons (91 within- and between-network connectivities and 247 network-subcortical connectivities).

### Total problems and network connectivity

Similarly, the effect of total problems on 338 unique network connectivities were tested. Thirteen associations between total problems and network connectivity survived FDR correction (see Fig. 2). Interestingly, the same three network connectivities impacted by total sleep disturbance were also influenced by total problems, i.e. DMN-DAN (*β* = 0.00033, *R*^*2*^ = 0.0655, *p* < 1e-7), DMN-DMN (*β* = -0.00026, *R*^*2*^ = 0.0497, *p* = 1.4e-4), and DAN-DAN (*β* = -0.00032, *R*^*2*^ = 0.0245, *p* = 1.6e-5).

**Figure 2.**
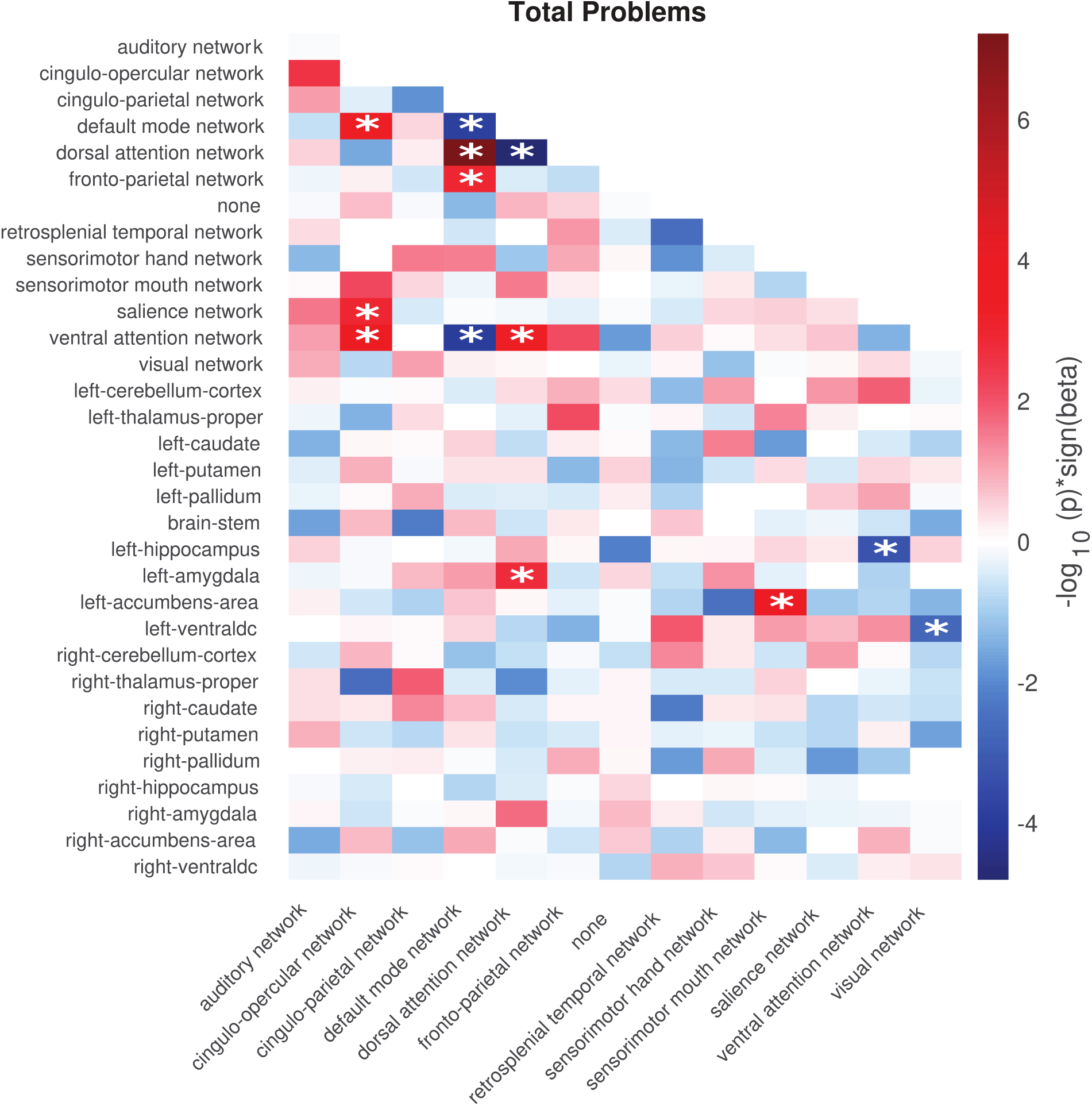
The effect of total problems on network connectivity measures. Color represents negative log-transformed *p* value times the sign of beta value of total problems. That is, red/blue means positive/negative association between total problems and network connectivity measures, respectively. Only unique network connectivity measures were shown in the matrix (i.e. top right was intentionally left blank). * denote network connectivity that survived FDR correction for multiple comparisons out of 338 unique comparisons (91 within- and between-network connectivities and 247 network-subcortical connectivities).

### Mediation analysis

Given that the impact of TSD and TP converged on the same three network connectivities (DMN-DAN, DMN-DMN, DAN-DAN), we further conducted a mediation analysis to explore the underlying mechanism by which TSD influences TP, or TP influences TSD, through a common mediator variable (DMN-DAN, DMN-DMN, or DAN-DAN connectivity). In other words, we tested whether these three network connectivities mediated the relationship between total sleep disturbance and total problems. We found that all three network connectivity measures significantly mediated the effect of sleep disturbance on total problems (all *p* < 0.005; 95% CI did not include 0, see Fig. 3 A-C). However, only the within-network connectivity (DMN-DMN) significantly mediated the effect of total problems on sleep disturbance (*p*=0.016; *β* = 0.0007; 95% CI, 0.0001-0.0014, see Fig. 3 D).

**Figure 3.**
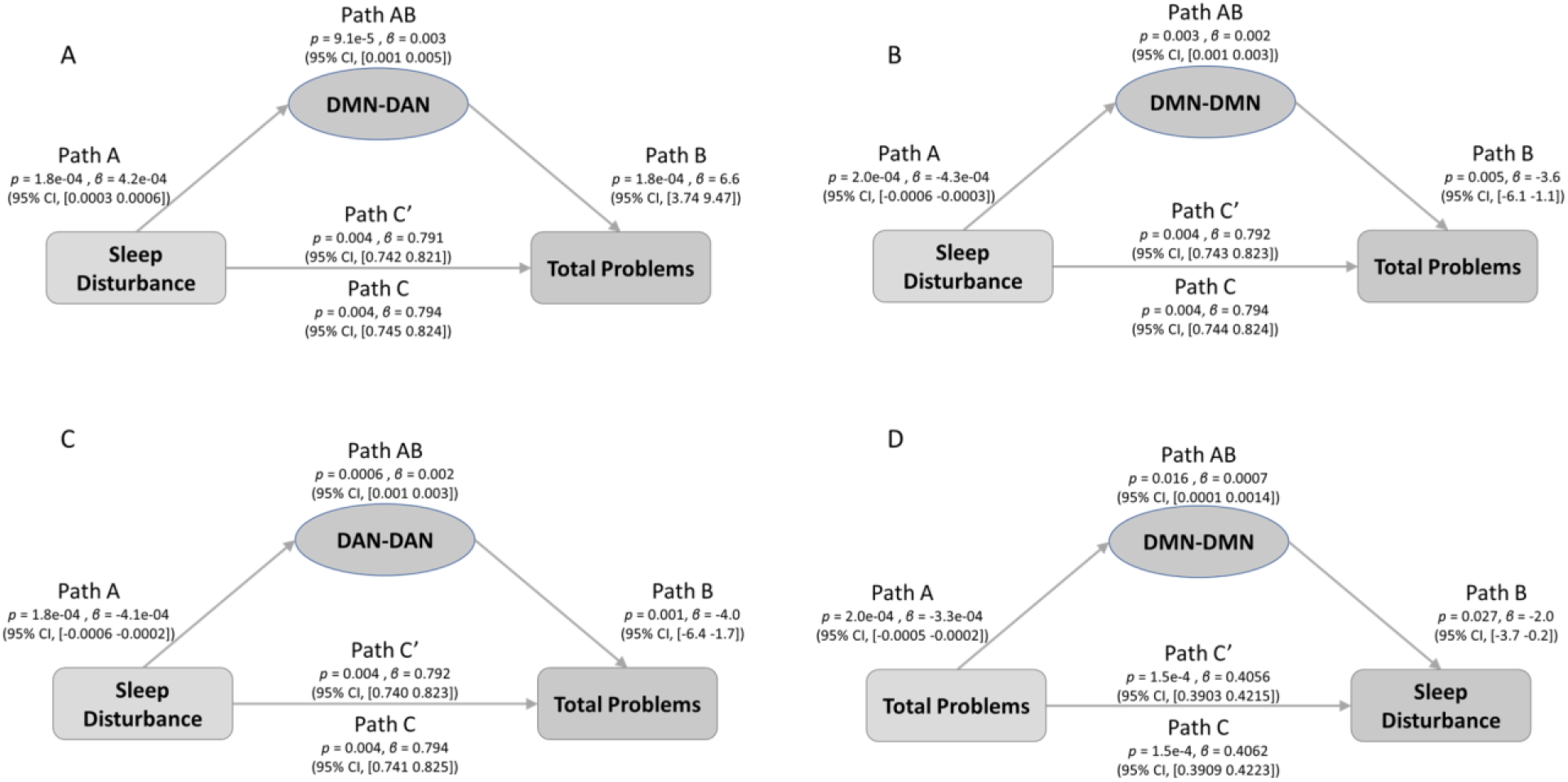
Network connectivities mediated the relationship between total sleep disturbance and total problems (A-C) and the relationship between total problems and total sleep disturbance (D). ***A***, DMN-DAN mediated the effect of total sleep disturbance on total problems (Path AB: *p*=9.1e-5; *β* = 0.003; 95% CI, 0.001-0.005). ***B***, DMN-DMN mediated the effect of total sleep disturbance on total problems (Path AB: *p*=0.003; *β* = 0.002; 95% CI, 0.001-0.003). ***C***, DAN-DAN mediated the effect of total sleep disturbance on total problems (Path AB: *p*=0.0006; *β* = 0.002; 95% CI, 0.001-0.003). ***D***, DMN-DMN mediated the effect of total problems on total sleep disturbance (Path AB: *p*=0.016; *β* = 0.0007; 95% CI, 0.0001-0.0014). DMN, default mode network; DAN, dorsal attention network.

### Correlation analysis

Follow-up partial correlation analysis showed that DMN-DAN and DAN-DAN were significantly correlated with the Total Cognitive Composite Fully-Corrected T-score (*r* = -0.0361 and 0.0231, *p* = 0.0008 and 0.032, FDR corrected *p* = 0.0025 and 0.0483, respectively), suggesting that these two network connectivities might be important for general cognitive functions in preadolescence.

### Longitudinal mediation analysis

Longitudinal mediation analysis revealed that all three network connectivities mediated the effect of total sleep disturbance at baseline on total problems one year later, even after controlling for baseline total problems (all *p* < 0.005; 95% CI did not include 0, see Fig. 4 A-C). Interestingly, distinct from the pattern that only DMN-DMN mediated the effect of total problems on total sleep disturbance at baseline, all three network connectivities mediated the effect of total problems at baseline on total sleep disturbance one year later, after adjusting for baseline total sleep disturbance (all *p* < 0.005; 95% CI did not include 0, see Fig. 4 D-E).

**Figure 4.**
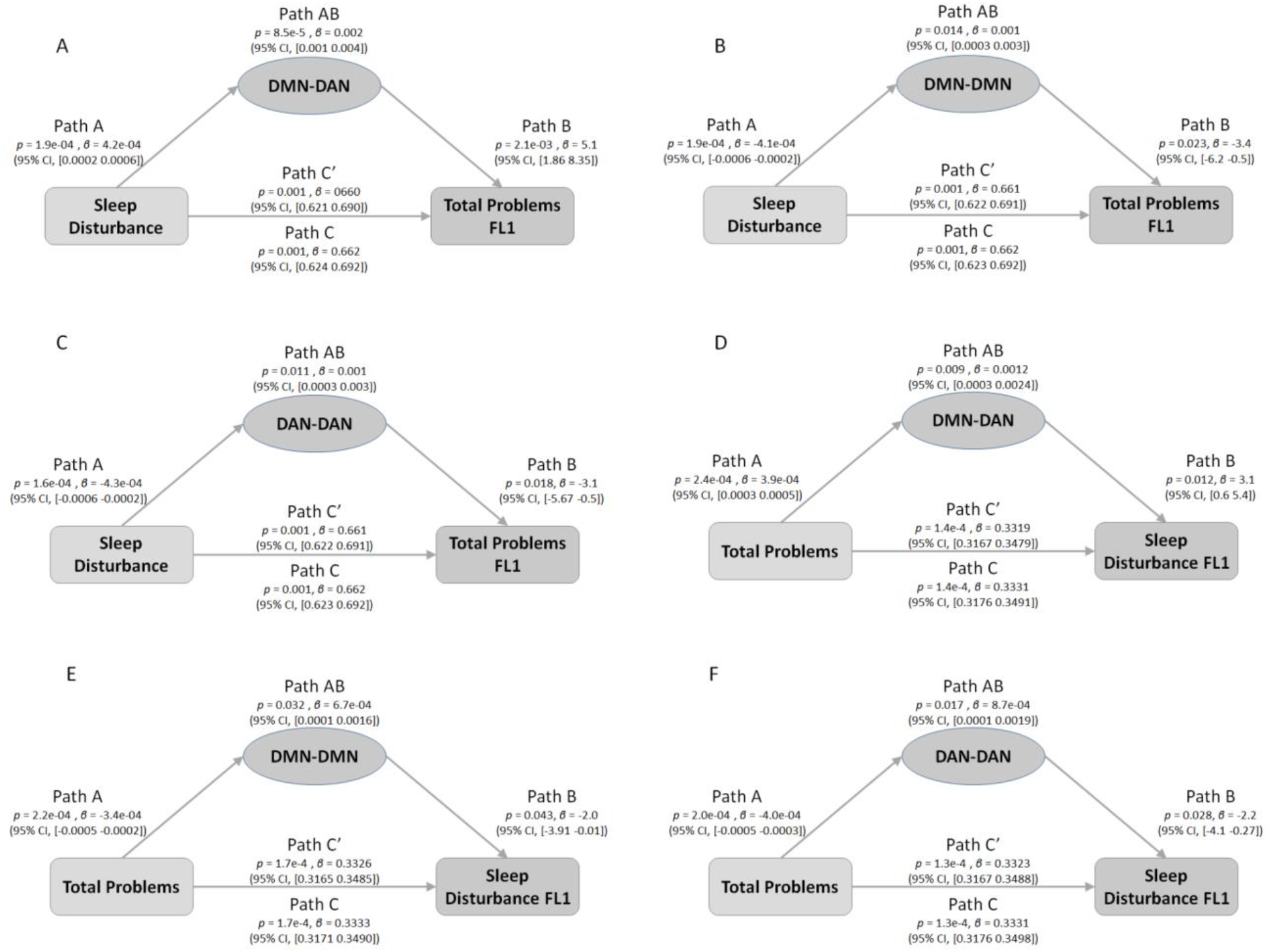
Network connectivities mediated both the relationship between total sleep disturbance (at baseline) and total problems (at one year follow-up, FL1) (A-C) and the relationship between total problems (at baseline) and total sleep disturbance (at FL1) (D-F). ***A***, DMN-DAN mediated the effect of total sleep disturbance at baseline on total problems at FL1 (Path AB: *p*=8.5e-5; *β* = 0.002; 95% CI, 0.001-0.004). ***B***, DMN-DMN mediated the effect of total sleep disturbance at baseline on total problems at FL1 (Path AB: *p*=0.014; *β* = 0.001; 95% CI, 0.001-0.003). ***C***, DAN-DAN mediated the effect of total sleep disturbance at baseline on total problems at FL1 (Path AB: *p*=0.011; *β* = 0.001; 95% CI, 0.001-0.003). ***D***, DMN-DAN mediated the effect of total problems at baseline on total sleep disturbance at FL1 (Path AB: *p*=0.009; *β* = 0.0012; 95% CI, 0.0003-0.0024). ***E***, DMN-DMN mediated the effect of total problems at baseline on total sleep disturbance FL1 (Path AB: *p*=0.032; *β* = 6.7e-4; 95% CI, 0.0001-0.0016). ***F***, DAN-DAN mediated the effect of total problems at baseline on total sleep disturbance at FL1 (Path AB: *p*=0.017; *β* = 8.7e-4; 95% CI, 0.0001-0.0019). Each longitudinal mediation analysis controlled for the corresponding baseline data of the dependent variable, e.g., for diagram A, baseline total problems was added as a covariate. DMN, default mode network; DAN, dorsal attention network; FL1, one year follow-up.

## Discussion

Based on data from a large cohort of preadolescents enrolled in the ABCD dataset study, our work identified robust relationships between network connectivity, cognition, sleep disturbance, and mental health. Total sleep disturbance and total problems both impact network connectivity and their relationship is mediated by network connectivity. The effects of sleep disturbance on total problems is mediated by three network connectivity measures, i.e. DMN-DMN, DMN-DAN, and DAN-DAN. In contrast, only DMN-DMN mediates the effect of total problems on sleep disturbance. Follow-up analyses showed that the degree to which DMN and DAN are anticorrelated (as two segregated modules) significantly correlates with the total cognition score. Longitudinal analysis revealed that total sleep disturbance and total problems at baseline can each predict the other one year later through the identified three network connectivities. These results confirm our hypothesis that sleep disturbance and mental disorders have similar impact on resting-state functional connectivity. Using a data-driven approach, we discovered that their impact converged in the between- and within-network connectivities in the DMN and one of its anti-correlated networks, DAN. Our findings suggest a common neural mechanism through which sleep disturbance and mental problems may exacerbate each other at both baseline and at one-year follow-up.

DMN is generally considered an integrated system that is associated with many different aspects of self-related mental processes such as autobiographical memory, internal thoughts, emotion regulations (Menon, 2011). Given these roles, it is not surprising that abnormal connectivity within DMN has been involved in almost every major psychiatric disorder, including dementia, schizophrenia, anxiety and depression, autism, and ADHD (Broyd et al., 2009). Consistent with these observations, we found that total problems, a summary score of various mental problems, was negatively correlated with DMN’s within-network connectivity (DMN-DMN) in the current study. That is, higher scores in total problems were correlated with weaker connectivities between brain regions within DMN. Moreover, we demonstrated that total problems had an indirect detrimental effect on sleep disturbance through DMN-DMN in preadolescence.

Unlike DMN, DAN is considered a “task-positive” network (Fox et al., 2005). Under externally directed cognitive tasks (e.g., visual search), DAN activation and DMN deactivation usually co-occur. This anticorrelation between DMN and DAN emerges in childhood and continues to develop during adolescence (Fair et al., 2009). In the current study, we found that both sleep disturbance and total problems were associated with the connectivity strength between DMN and DAN. Moreover, we found that a less separated DMN-DAN is also correlated with lower total cognition scores, suggesting that a segregation between DMN and DAN might be a critical factor accounting for one’s cognitive performance, in line with a previous study showing that variations in network configurations, e.g. less segregation of DMN-DAN affects the development of cognition (Gu et al., 2015).

Emerging evidence suggests that symptom severity of various mental disorders and sleep disturbance could impact each other in a bi-directional manner (Tesler et al., 2013). For example, less sleep disturbance was found in children who received treatment for anxiety disorders compared to those who received placebo (Alfano et al., 2007). In youth with mood disorders, sleep disturbance was associated with more severe symptomatology, longer episodes, and increased risk for relapse (Emslie et al., 2012; Liu et al., 2007). To our knowledge, the current study first explains the co-occurrence and bi-directionality of mental disorders and sleep disturbance by identifying a shared network mechanism between sleep disturbance and total problems, i.e. within- and between-network connectivity in DMN and DAN. Mediation analysis and longitudinal mediation analysis further confirmed that sleep disturbance and mental disorders could affect each other via these three network connectivities (DMN-DAN, DAN-DAN, and DMN-DMN).

The specificity of the functional networks (DMN-DAN, DAN-DAN, and DMN-DMN) as the mediator identified throughout longitudinal mediation analyses can further inform the design and application of real-time fMRI connectivity-based neurofeedback training. Real-time fMRI neurofeedback (rt-fMRI-NF) is an emerging non-invasive technique to modulate aberrant neurocircuitry, with the potential to induce long-term symptom reductions in patients with various mental disorders (Linhartová et al., 2019). In our pathway model, the DMN-DAN connectivity mediated both the bidirectional and longitudinal relationship between mental disorders and sleep disturbance. Thus, regulating DMN-DAN connectivity strength can potentially result in reduced mental problems and sleep disturbance, potentially yielding a positive and long-term effect in individuals at-risk for mental disorders and sleep disturbance.

Several limitations in the current study should be noted. First, the effect sizes found in the current study with large sample sizes are relatively small compared to those reported in the literature based on much smaller sample sizes. Despite a small r value of the correlation between DMN-DAN and total cognition (−0.036), according to ABCD analysis guidelines (Dick et al., 2021), a -0.036 r value in about 10,000 participants has statistical power higher than 0.9, which is considered well-powered. In addition, a previous large sample-size study found even the most significant results can only explain around 1% of the total variances (Miller et al., 2016). Second, we focused on the high-level commonality between sleep disturbance and various mental disorders (rather than each type of mental disorder). Thus, we did not investigate the relationship between sleep disturbance and each specific type of mental disorder. Caveat is needed when interpreting the current findings with regard to specific mental disorders, e.g. depression, anxiety disorders, etc.

Taken together, our results shed light on a shared and stable neural mechanism between sleep disturbance and mental disorders. The network connectivity between DMN and DAN that mediated the bidirectional and long-term relationship between sleep disturbance and mental disorders are key target brain networks for neurofeedback training or behavioral therapy. Future work may examine the effectiveness of regulating connectivity strength between DMN and DAN in inducing positive behavioral change related to mental health and sleep problems.

## Data Availability

Data were provided from the Adolescent Brain Cognitive Development (ABCD, https://abcdstudy.org) study. Codes were available from the corresponding author upon reasonable requests

## Acknowledgement

This study was supported by NIH/NIA grants R01AG060054, R01 AG070227.

